# Synonym Augmentation for Rare Disease Identification in Unstructured Data

**DOI:** 10.64898/2026.05.11.26352910

**Authors:** Jaber Valinejad, Sungrim Moon, Yanji Xu, Qian Zhu

## Abstract

The significant challenges associated with rare diseases in the medical and research domains include the scarcity of information, which is often confined to unstructured formats. Although existing approaches provide valuable insights, there is a need to develop effective methods to identify information pertinent to rare diseases for advancing rare disease research. We identified mentions of rare diseases in relevant texts and assessed their relevance using derived scores, the confidence score and semantic similarity from a fine-tuned BioMedBERT encoder. This encoder was fine-tuned using rare disease related text from Online Mendelian Inheritance in Man (OMIM), Orphanet, a manually validated dataset, and STS benchmark datasets. The process of identifying meaningful rare disease mentioned was presented through two case studies that retrieved relevant NIH-funded projects, utilizing a generated knowledge graph in Neo4j to host data on 2,067 GARD diseases with over 320,000 NIH funded projects. Through various case studies with NIH-funded projects related to rare diseases, we demonstrated the effectiveness of our approach in systematically providing rare disease related data to enhance our understanding of rare diseases for future investigations.

## 1. Introduction

Rare diseases refer to those affecting fewer than 200,000 individuals each in the United States. For the extremely rare diseases, there might be only very few people affected. For instance, Progeria, or Hutchinson-Gilford progeria syndrome (HGPS) can be noticed in 1 out of 20 million people [1]. These conditions are often poorly understood, with unclear biological mechanisms and a lack of effective treatments, leading to diagnostic delays and ongoing high medical costs [2] [3]. Despite the advancements in NLP techniques, challenges persist in integrating domain-specific knowledge and standardizing terminology from unstructured data [4].

In recent years, various research endeavors have aimed to gather and extract information relevant to rare disease by employing Natural Language Processing (NLP) and Large Language Models (LLM). He, D., et al. systematically analyzed the existing literature on the application of AI in the treatment of rare diseases, providing a comprehensive overview of the current state of research [5]. Martínez-deMiguel, C., et al. developed RareDis, a gold standard corpus annotated with rare diseases and their clinical manifestations, to extract relevant information about rare diseases [6]. Taboada, M., et al. demonstrated the automatic identification and semantic annotation of relevant patient case reports to capture the phenotype of cerebrotendinous xanthomatosis, a rare condition that affects the body’s ability to metabolize fats known as cholesterols [7]. Shen, F., et al. applied association rule mining algorithms on electronic medical records (EMR) to extract significant phenotype-disease associations, enriching existing rare disease resources such as the Human Phenotype Ontology (HPO) and Orphanet, and generating phenotype-disease bipartite graphs [8]. Additional effort was made in using deep learning models for rare disease relevant information extraction. Segura-Bedmar et al. explored bidirectional Long Short-Term Memory (BiLSTM) networks and deep contextualized word representations based on Bidirectional Encoder Representations from Transformers (BERT) to recognize rare diseases and their clinical manifestations [9]. Dong, H., et al. investigated recent advancements in NLP for identifying rare diseases from clinical notes using ontologies and weak supervision-based BERT models [10]. Shyr, C., et al. identified and extracted rare disease phenotypes using prompt learning with LLMs in zero- and few-shot settings [11]. Pokhrel [12], K., et al. developed OrphaGPT, a customized user interface powered by a fine-tuned GPT-3.5 Turbo model, enabling users to engage in detailed conversations about rare diseases [12]. OrphaGPT can also be used for rare disease identification. Wang, A., et al. fine-tuned Llama 2 (Llama2-7B) for rare disease concept normalization using standardized HPO names and selected synonyms with their identifiers [13].

These studies illustrate a range of challenges in the field of rare disease recognition and extraction using NLP and LLM techniques. Many approaches heavily rely on existing ontologies, such as Unified Medical Language System [14](UMLS) and Orphanet database [15], which, while powerful, may lack comprehensive coverage, particularly for very rare diseases, leading to potential missed cases or incorrect matches. Additionally, the generalizability of models trained on specific datasets, like MIMIC-III, is questioned, as these models may not perform well across different clinical settings or datasets due to variations in medical language and note structures. The use of advanced models like BERT and ChatGPT for tasks such as few-shot learning presents further challenges, including model sensitivity to training examples, risk of overfitting, and computational complexity. Moreover, the limited availability of annotated datasets, as seen in the RareDis corpus, poses significant hurdles in training robust models. The integration of LLMs like OrphaGPT with scarce datasets remains underexplored, highlighting the need for more research in this area. Importantly, none of the studies explicitly determine whether the focus of any given text or unstructured dataset is on a specific rare disease, and could limit the precision and relevance of the extracted information. Lastly, the reliance on template-based approaches for phenotype identification, as seen with the HPO, may limit the depth and flexibility of these methods, potentially restricting their applicability in more complex scenarios. To fulfill the gaps and limitations of identifying rare disease specific information from the aforementioned studies, we extended our previous work [4] [16] with a two-step process in obtaining rare disease related information from NIH grant funding data. To ascertain the relevance of a given unstructured textual data to a rare disease, we predefined two scores, confidence score and semantic similarity to assess the focus of the given text is on the rare diseases. The confidence score is used along with negation detection and sentence tense checking, while the semantic similarity score is fine-tuned BiomedBERT. We described the resources used in this study and each step of our process in the Materials and Methods section, corresponding results are summarized in the results section. Three case studies were performed to demonstrate the use of advancing rare disease research with identified research projects.

## 2. Materials and Methods

In this study, we aimed to develop an automated workflow for identifying rare disease-related information, exemplified by a case study on retrieving relevant NIH-funded projects. The workflow comprised four main components: rare disease data preparation, project data preparation, potential rare disease-based project identification, text relevance assessment, including semantic similarity score calculation, and confidence score measurement. Each of these components is described in detail in the sections below. Figure 1 outlines the pipeline for identifying rare disease related NIH-funded projects.

**Figure 1.**
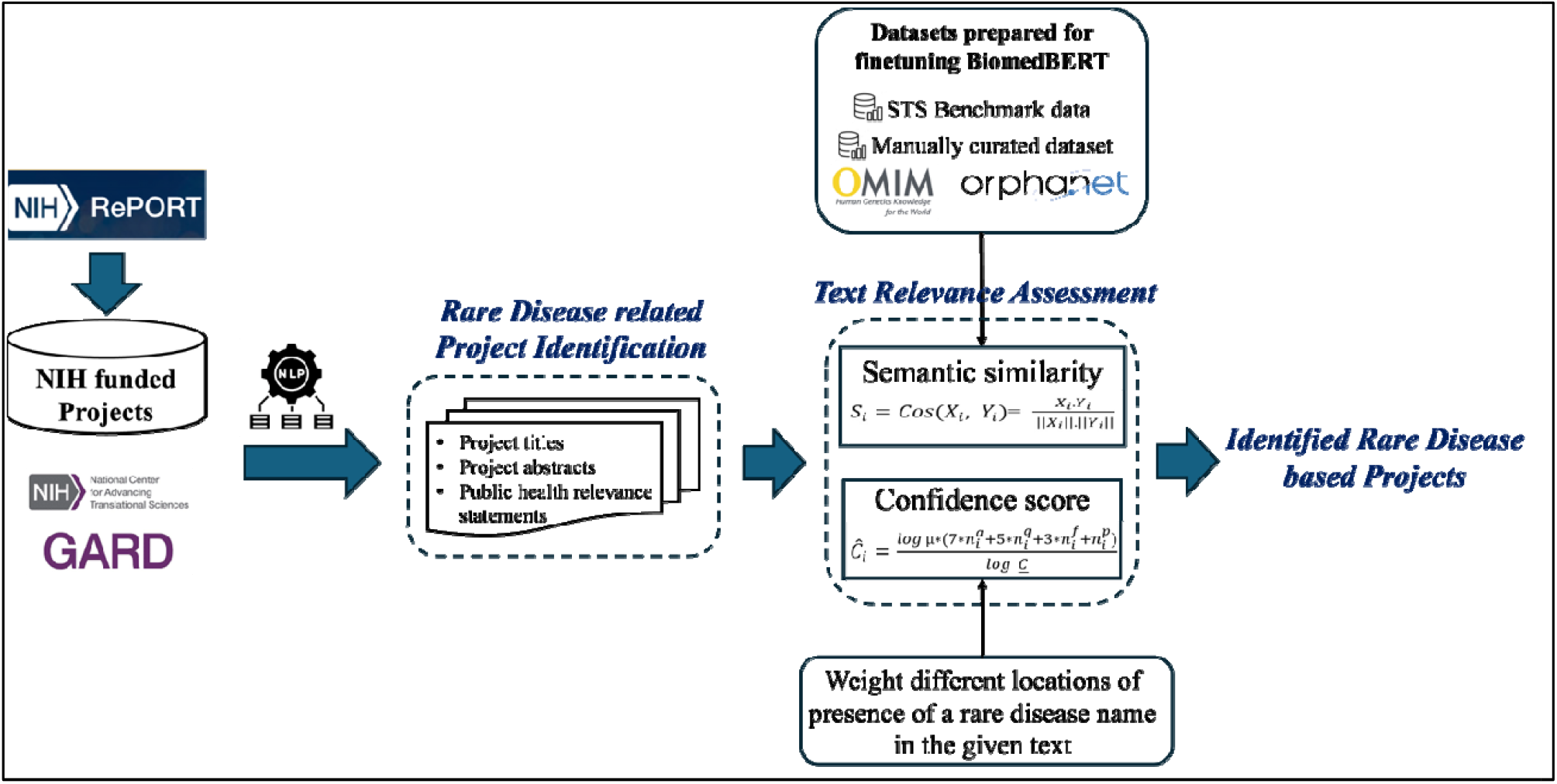
Rare Disease based text identification with an example of NIH Funded Projects.

### 2.1. Rare Disease Data Preparation

We obtained rare disease data from Genetic and Rare Diseases Information Center (GARD) [17], a program dedicated to rare diseases and governed by the National Center for Advanced Translational Science (NCATS)/NIH, including rare disease IDs, rare disease names, their synonyms and other rare disease related information [18]. To prepare these rare disease names and their synonyms from GARD for the task of project mapping, we normalized them via four steps, 1) excluding terms containing the phras “susceptibility to,” as these do not align with the definition of rare diseases^1^, 2) employing text stemming [19] to normalize terms in their basic forms, 3) implementing the bag-of-words (BOW) [20] when the word count is less than three^2^, 4) Retaining synonyms longer than five letters that are unlikely to be abbreviations^3^.

### 2.2. Rare Disease Project Preparation

The NIH grant data sourced from NIH RePORTER [21] is a repository of grant administrative information, including NIH funded project application ID, fiscal year for funding, project terms, project abstract, project title, public health relevance statement, and other relevant details. We leveraged the NIH ExPORTER API [22] to access and retrieve the NIH grant data tailored to our specific use case. To ensure the target statements contained pertinent details about the funded project aims with a research focus on rare diseases, we analyzed the project title, abstract, and public health relevance statement if applicable from each project. In addition, we excluded sentences written in the past tense, which is most likely describing previous work or background information, instead of the primary focus of the project. We performed negation checking with spaCy [23] on each sentence to exclude those containing negative representations from further consideration. Furthermore, we implemented text stemming to normalize words into their root forms.

### 2.3. Rare Disease Related Project Identification

We focused on identifying mentions of a rare disease or their synonyms within the specific section of the given text, from project titles, public health relevance statements, and project abstracts. We assumed that a rare disease mentioned in the project title, is most likely the primary research focus of the project. Hence, the mapping process halts if any rare disease was found in the title. Otherwise, it continued with the public health relevance statement. If none were identified, the algorithm then analyzed the abstract to detect any mentions of rare diseases if applicable.

From each given piece of information from project titles, public health relevance statements, and project abstracts, we identified any mentions of rare diseases based on exact string match. Furthermore, we applied stemming to the texts and disease names/synonyms for additional mappings. For example, ‘Scheie syndrome’ (GARD:0012561) was matched to a project with a project application ID of “3152429” based on mapping with the project title ‘therapy of a model of hurler/scheie syndrome’. By applying stemming to the project title, it was matched to a more accurate rare disease, ‘Hurler-Scheie syndrome’ (GARD:0012560). When multiple instances of rare diseases were present in the text, we summed the number of mentions for each rare disease. A greater number of mentions of a rare disease might indicate a higher level of relevance, thus favor those with a higher frequency of mention. When multiple GARD names are found, we prioritize the longer name if its number of mentions equals or greater than others. In this case, ‘Hurler-Scheie syndrome’ (GARD:0012560) was retained for the above example.

### 2.4. Text Relevance Assessment

#### 2.4.1. Semantic Similarity Score

Although our mapping approach can successfully identify mentions of rare diseases in the text, it needs to further determine whether the text is scientifically relevant to the identified rare disease. We designed the semantic similarity score to ascertain the relevance by considering the context surrounding the mention of rare diseases in the text. The semantic similarity score, ranging from 0 to 1, signifies the likelihood that the project is relevant to the identified rare disease, with higher scores indicating a higher relevance. We employed BiomedBERT[24], pre-trained using abstracts from PubMed and full-text articles from PubMedCentral for biomedical text embedding. Since BiomedBERT is an encoder model, it is necessary to fine-tune it for the task of semantic similarity calculation. Thus, we created six datasets for the fine-tuning task. The six sets can be accessed from the supplemental files named FinetunningDataset.xlsx, which contains six sheets corresponding to six datasets.

##### 2.4.1.1. Data Curation

###### Set 1

Manually curated dataset. To prepare a labeled dataset for finetuning the BiomedBERT model, we randomly selected 10,000 NIH funded projects mapped to 1,989 rare diseases from the step 2.3. Four biomedical scientists, as co-authors holding PhD degrees, manually reviewed those mappings. To ensure their review conducted consistently aligned with the study requirements, we created manual curation guidelines (please see the supplemental file named ManualCurationGuidelines.docx) with detailed descriptions of the inclusion/exclusion criteria for labeling each mapping between rare diseases and projects as relevant vs irrelevant. By the end, the reviewers labeled those projects as “Relevant” if their research focus was on the identified rare disease, otherwise labeled as “Irrelevant”. Any further comments/notes towards their decisions were captured for further discussion during their review.

###### Set 2

The Semantic Textual Similarity(STS) Benchmark dataset [25], consisting of 8,628 sentence pairs was selected. Although the STS dataset was primarily developed and used in general domains, it can be effectively applied on those available sentence pairs, shown in Table 1 for fine-tuning a transformer model for semantic similarity computing [26]. The similarity scores in the dataset range from 0 to 5, which was normalized to a range of 0 to 1 in this study.

**Table 1.** Examples from the STS Benchmark dataset.

| Sentence 1 | Sentence 2 | Similarity score |
| --- | --- | --- |
| A plane is taking off. | An airplane is taking off. | 5 |
| A man is mowing a lawn. | A woman is cutting a lemon. | 0.2 |
| A woman is dancing. | A man is talking. | 0 |

###### Set 3

A dataset with definitions of rare diseases from OMIM [27] and Orphanet [15]. It comprises 2,335 disease definition pairs with the semantic similarity predefined as 1.

###### Set 4

A dataset with GARD disease names and their disease definitions. It comprises 8,419 pairs of disease name and its definition with the semantic similarity as 1.

###### Set 5

A dataset with GARD disease names and randomly paired with non-medical sentences from STS Benchmark. It comprises 8,419 pairs of disease name and non-medical sentences with the semantic similarity as 0.

###### Set 6

A dataset with GARD disease synonyms and their disease definitions. It comprises 28,030 pairs of disease synonym and its definition with the semantic similarity as 1.

##### 2.4.1.2. Model Development

BiomedBERT uses multiple transformer layers to process the embeddings. Each layer consists of self-attention to capture long-range dependencies and feed-forward neural network components. BiomedBERT outputs token embeddings for each token in the input sequence. We then applied the pooling layer to aggregate these token embeddings into a single fixed-size sentence embedding. Mean pooling computes the average of all token embeddings, as shown in Eq. 1.

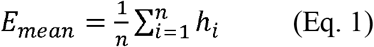

Where H=[*h*_1_, *h*_2,_ …., *h*_*n*_] is the sequence of token embeddings generated by the transformer model for a given input sentence, where h_i_ is the embedding for the i-th token. Next, we used cosine similarity to find the similarity between two embeddings. For a sentence pair <X, Y>, BiomedBERT is to calculate a similarity score between X and Y using Eq. 2.

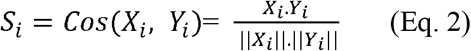

Where S_i_ is a semantic similarity score for a sentence pair <X, Y>. The index i corresponds to the index of the NIH funding project. To fine-tune the model, we iterated over Sets 1-6 in batches, calculated the loss for each batch, and updated the model parameters using the AdamW optimizer.

Eq. 2 is used to calculate the similarity score for both training and test input embeddings. The predicted similarity score for the i-th project was calculated by Eq. 3 where the training input embeddings are used for fine-tuning purposes.

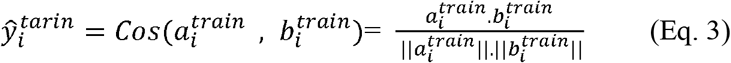

Where the input embeddings are 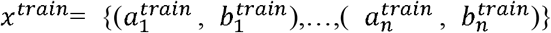 and the predicted semantic similarity is 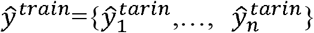. The superscript ‘train’ indicates that the input embeddings correspond to the training datasets from Sets 1-6. We employed mean square error as the loss function, *L*_*batch*_, for each batch size B using Eq. 4 and then update the model parameters using Eq. 5.

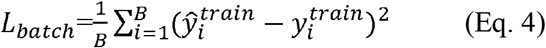

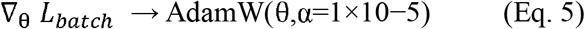

Where the true semantic similarity is 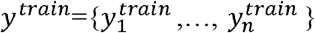. Additionally, θ represents the model parameters, and α is the learning rate.

##### 2.4.1.3. Semantic Similarity Evaluation

The semantic similarity score ranges between 0 and 1, and different thresholds indicates the level of scientific relevance of the project to the identified rare disease. For each threshold, we calculated the False Positives (FP), True Positives (TP), False Negatives (FN), True Negatives (TN), and accuracy to determine the most appropriate threshold. The accuracy is calculated by using Eq. 6.

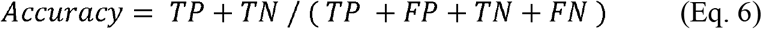

We also used Eqs. 7-10 to calculate FP Percentage, Precision, Recall, F1-score:

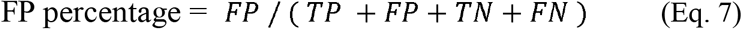

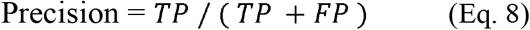

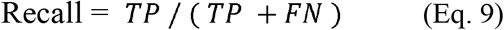

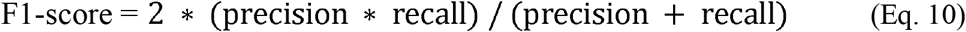

#### 2.4.2. Confidence Score

We assumed that the presence of a rare disease name in different locations of the given text signifies the level of relevance of the rare disease as the primary research focus of the project. Consequently we defined a confidence score as a weight to the different places where the disease was mentioned and assigned the score to the mappings:

1. If the GARD name or its synonyms are found in the title, we assigned a weight of 20.
2. If the GARD name or its synonyms are found in the abstract:
  - If it is in a sentence with “the goal of” or ‘aim’, we assigned a weight of 7 for it.
  - If it is in the first sentence, we assigned a weight of 5 for it.
  - If it is in a sentence with future tense, we assigned a weight of 3 for it.
  - If it is in a sentence with present tense, we assigned a weight of 1 for it. In summary, we computed the confidence score by calculating the sum of the frequency of mentions with the predefined weights using Eq. 11.

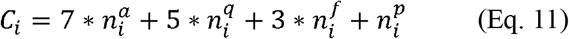

where *C*_*i*_ represents the non-normalized confidence score for the mapping with the rare disease name found in the project i; 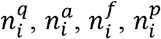 denote the number of occurrences of rare disease names in the first sentence, sentences containing ‘aim’ or ‘the goal of’, future tense sentences, and present tense sentences respectively.
3. If we find the GARD name or its synonyms in the public health relevance statement, we double the weights compared to those in the abstracts.

To normalize the confidence score, we used the Eq. 12:

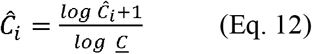

where *<u>C</u>* is a normalized confidence score. Similar to semantic similarity score, the normalized confidence score ranges between 0 and 1. Adding 1 to the numerator is to ensure that the normalized confidence score is 0 when there is no mention of a rare disease. Based on the normalized confidence score, we considered different thresholds to determine the level of scientific relevance of the project to the identified rare disease. So, the normalized confidence score is obtained using Eq. 13.

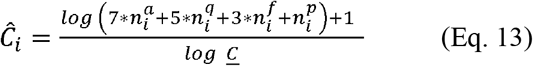

### 2.5. NIH Grant Funding Knowledge Graph Development

In this study, we analyzed the entire NIH grant funding data, and subsequently developed a graph database hosted in a neo4j to maintain the analyzed data, which can be freely accessed via the database named rdas_gfkg at https://rdas.ncats.nih.gov/browser/.

## 3. Results

### 3.1. Results for Rare Disease Data Preparation

We collected a total of 12,004 rare disease entries from GARD. After excluding 138 entries containing the phrase “susceptibility to,” 11,866 rare diseases and 14,423 associated synonyms were left. Of these rare diseases/synonyms, 5,336 terms composed of two words, were processed using the Bag of Words (BOW) technique. Next, we excluded 1,224 terms with fewer than five characters, resulting in 30,401 terms remaining. Applying stemming to the rare diseases—excluding those with the phrase ‘susceptibility to’— and to their synonyms, and using BOW on the stemmed terms while removing those with fewer than five characters, generated an additional 30,239 new terms. As a result, a total of 60,640 terms were applied to map to the NIH projects. Table 2 summarizes the preparation of the rare disease data.

**Table 2.** Results for rare disease data preparation.

|  | Rare diseases | Containing the phrase "susceptibility to" | Synonyms | Two words (for BOW) | Fewer than five characters | Total |
| --- | --- | --- | --- | --- | --- | --- |
| **Exact match** | 12,004 | 138 | 14,423 | 5,336 | 1,224 | 30,401 |
| **Stemming** | 12,004 | 138 | 14,303 | 5,337 | 1,267 | 30,239 |

### 3.2. Results of finetuning of BiomedBERT using Sets 1-6

*Sets 1-6* consists of 40,261 pairs of sentences and their associated semantic similarity. Our training dataset comprised 26,982 unique samples, with 6,433 for testing and 6,846 for validation. Model evaluation was conducted on the validation dataset after each epoch to track performance. Figure 2 shows the evaluation results for each epoch. The results show that the model is learning and generalizing effectively, with a steady decrease in training loss indicating successful error minimization. The validation loss also improves, though the rate of decrease slows as the model converges. The stability of the validation loss towards the end suggests the model is nearing optimal performance, and there is no significant overfitting, as both training and validation losses remain closely aligned.

**Figure 2.**
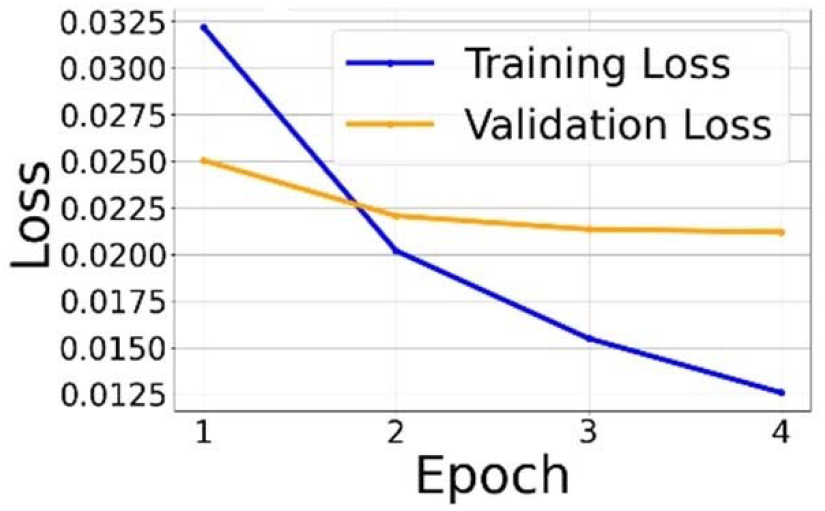
BiomedBERT training progress and performance metrics across epochs.

### 3.3. Evaluation Results on Rare Disease Relevant Project Identification

We utilized Set 1, which includes 1,822 NIH-funded projects labeled as either Relevant or Irrelevant, to evaluate our algorithm for rare disease relevant project identification. Figure 3 displays the distribution of average confidence scores and semantic similarity scores across two categories: Relevant and Irrelevant. The average normalized confidence score for projects labeled as Relevant is 0.83, compared to 0.56 for projects labeled as Irrelevant. Similarly, the average normalized semantic similarity score for project labeled as Relevant is 0.81, whereas projects labeled as Irrelevant is 0.5. While the confidence or semantic similarity scores increasing, the probability of a project being labeled as Relevant rather than Irrelevant increases, and vice versa.

**Figure 3.**
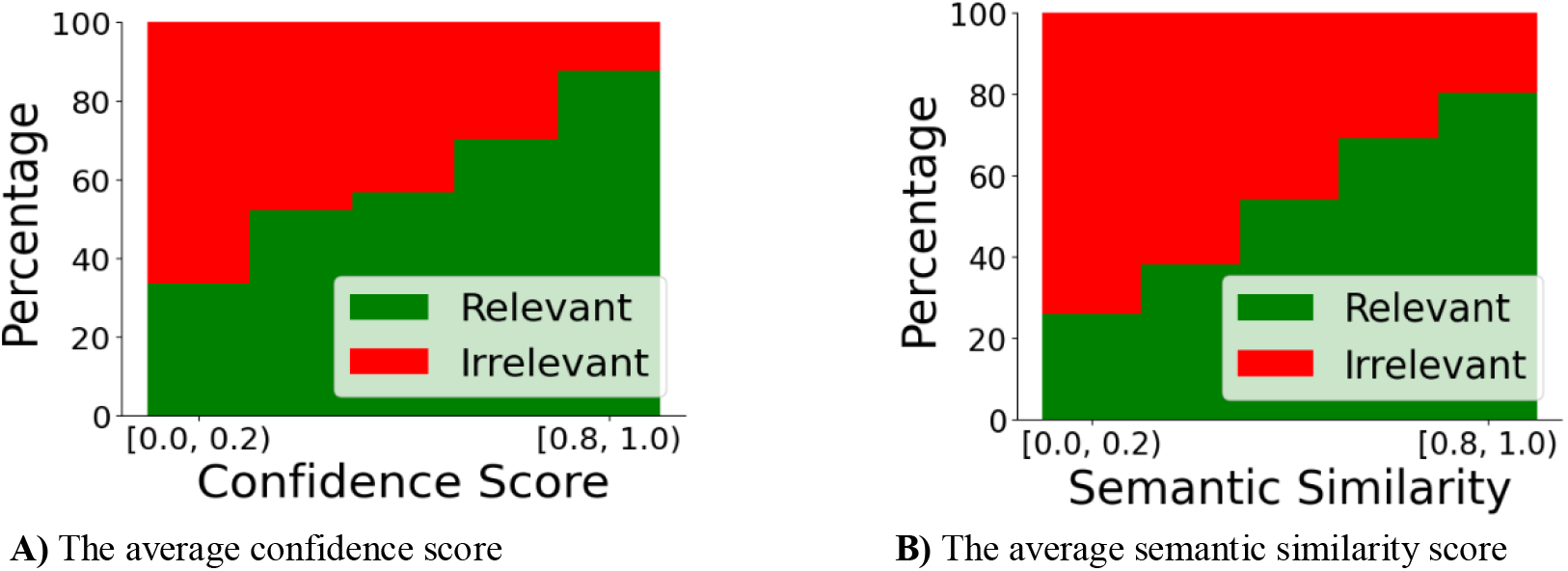
Evaluation results on rare disease identification using set 1 including 1822 NIH-funded projects. (A) The percentage of the average confidence score for two cases of Relevant and Irrelevant. (B) The percentage of the average semantic similarity score for two cases of Relevant and Irrelevant.

### 3.4. Results on NIH Grant Knowledge Graph

Figure 4 summarizes the statistical results within the data model we established for NIH grant knowledge graph. A total of 2,067 GARD diseases were associated with 320,120 projects, corresponding to 75,373 NIH-funded applications. Of these projects, 320,120 have titles, 145,991 have abstracts, and 135,342 include public health relevance statements. It’s important to note that a single project can be funded through multiple applications over several years, resulting in a higher count of mapped applications than unique project titles.

**Figure 4.**
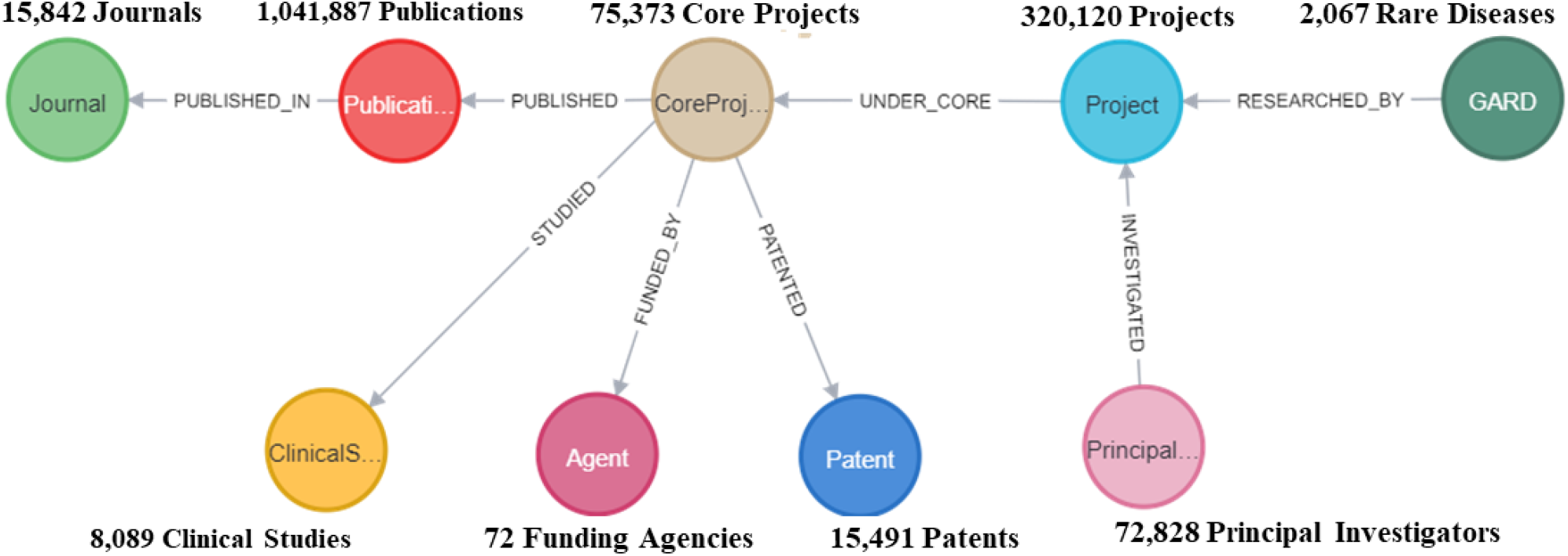
Statistical results of NIH grant knowledge graph in Neo4j.

## 4. Case studies

We conducted two case studies demonstrating how rare disease-related NIH grant funding data can be applied to identify research initiatives, address data limitations, and expedite medical breakthroughs in rare diseases. (1) assessing rare disease funding landscape to provide an overview of current funding trends, (2) analyzing the research landscape for Hemophilia B, and (3) identifying potential drugs for Primary Sclerosing Cholangitis (PSC). Cypher Queries, Neo4j’s graph query language, were developed to extract from the NIH grant knowledge graph for these case studies. These queries can be found in the supplementary File named CypherQueries.docx.

### 4.1. Case Study 1: Rare Disease Funding Landscape Analysis

We formulated Cypher Query 1 to obtain funding data by US states, as shown in Figure 5A. California, Massachusetts, and New York are the top three states that received more than $11 billion in funding for rare disease research. Obviously, the funding level is highly correlated (0.97) with the number of research papers, namely, more funding results in more publications.

**Figure 5.**
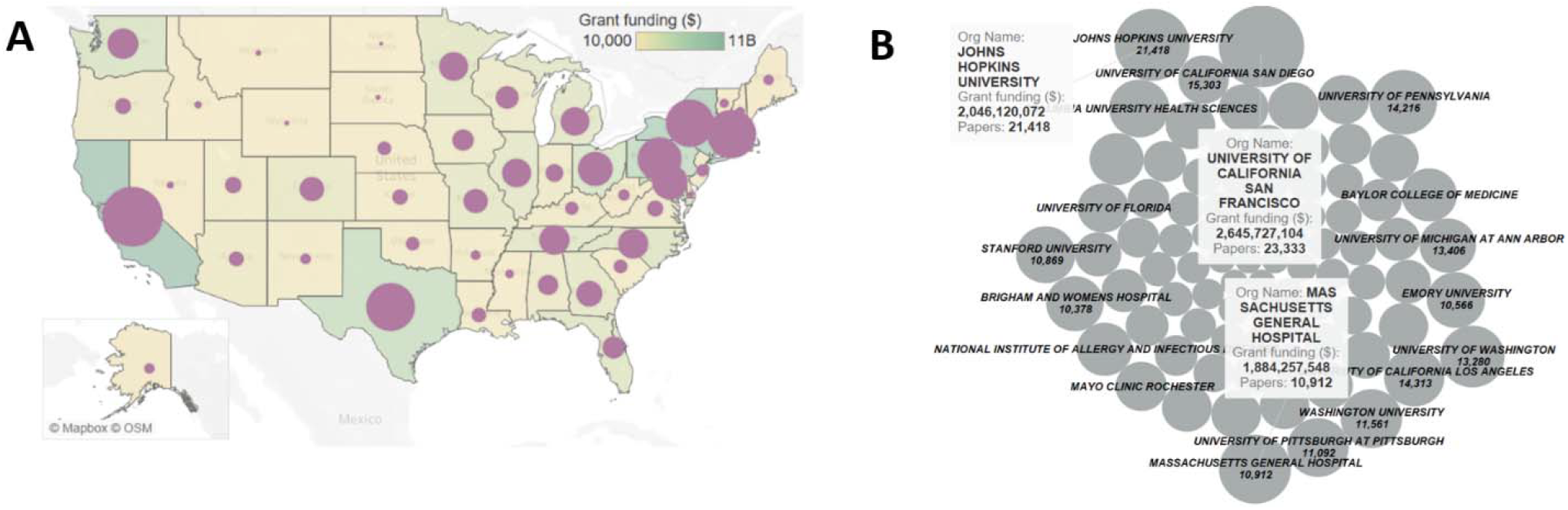
Funding landscape analysis. (A) The distribution of grant funding for rare disease research across U.S. states. Each state’s color indicates the funding level, and the circle size within each state represents the number of publications. (B) Grant funding distribution for rare disease research across U.S. institutions. The size of the circle within each state represents funding level.

#### Cypher Query 1

MATCH (n:Project)-[re:RESEARCHED_BY]-(g:GARD)

WHERE re.semantic_similarity > 0.6

MATCH (n)--(p:PrincipalInvestigator)

OPTIONAL MATCH (n)-[:UNDER_CORE]-(c:CoreProject)-[:PUBLISHED]-(e:Publication)

WITH p.org_state AS org_state, SUM(n.total_cost) AS total_cost1,

COUNT(DISTINCT e) AS num_papers

RETURN org_state, total_cost1, num_papers;

Composing Cypher Query 2, we extracted the funding data across U.S. research institutions, as depicted in in Figure 5B^1^, which highlights leading research universities and medical centers, such as the University of California San Francisco, Johns Hopkins University, and Massachusetts General Hospital, as major recipients of NIH funding for rare disease research. While it is generally expected that institutions receiving more funding will publish more research, this is not always the case. We can rank institutions based on the number of papers published per dollar of funding as a measure of research output. Among the institutions that received funding, Oregon Regional Primate Research Center, Mellon Pitt Corporation (MPC Corp), and Los Angeles County Harbor-UCLA Medical Center have the highest research outputs based on their funding (0.005802797, 0.005383557, 0.002069839 publications per dollar, respectively).

#### Cypher Query 2

MATCH (n:Project)-[re:RESEARCHED_BY]-(g:GARD)

WHERE re.semantic_similarity > 0.6

MATCH (n)--(p:PrincipalInvestigator)

OPTIONAL MATCH (n)-[:UNDER_CORE]-(c:CoreProject)-[:PUBLISHED]-(e:Publication)

WITH p.org_name AS org_name, SUM(n.total_cost) AS total_cost1,

COUNT(DISTINCT e) AS num_papers

RETURN org_name, total_cost1, num_papers;

### 4.2. Case Study 2: Research Landscape Analysis for Hemophilia B

Funded projects and publications show snapshots of their research goals and outcomes, which provides an opportunity to assess the current research status for those rare diseases systematically. We enumerated the assessment for Hemophilia B (GARD:0008732), a rare genetic disorder caused by a deficiency of Factor IX, which leads to improper blood clotting and prolonged bleeding [28]. According to FDA Orphan Drug Designations and Approvals, several approved drugs include generic names such as Coagulation Factor IX (recombinant) and etranacogene dezaparvovec-drlb, which were approved for treating hemophilia B [29].

With Cypher Query 3, we retrieved 2,301 projects and 151,848 associated publications. Subsequently we annotated the key biomedical terms from those project abstracts by exploring spaCy. Thus, we analyzed the funding data by stratifying with those annotated biomedical terms for Hemophilia B, as illustrated in Figure 6. Pie chart A illustrates the distribution of research conducted prior to 2000, while pie chart D highlights the distribution of more recent research from 2020 onwards.

**Figure 6.**
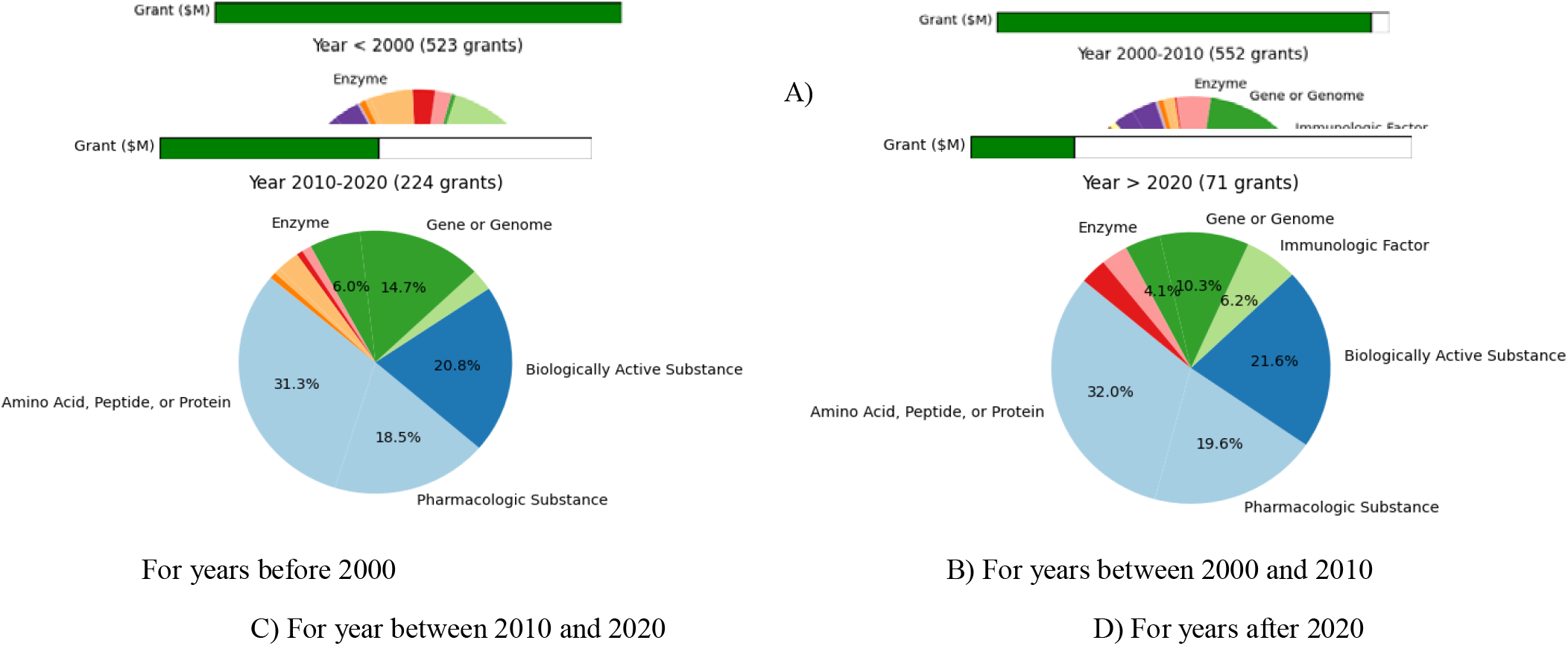
Distribution of annotated biomedical terms and grant funding allocated to Hemophilia B over time.

Before 2000, 523 grants totaling over $20 million were awarded for research across five designations and three approved treatments, laying the groundwork for future studies. Notable research focused on pharmacologic substances (103 projects), biologically active substances (124 projects), amino acids, peptides, proteins (195 projects), and enzymes (27 projects), enhancing the understanding of Hemophilia B mechanisms. From 2000 to 2010, 552 grants totaling over $19 million were allocated to research in one designation, expanding into gene or genome (49 projects) and immunologic factors (23 projects). Between 2010 and 2020, funding decreased, with 224 grants worth over $10 million awarded, yet gene or genome research interest increased from 9% to 14.7%. This period supported six designations and led to four approved treatments. After 2020, 71 grants totaling over $4.6 million resulted in five approved treatments, continuing the focus on Hemophilia B, although with lower funding levels.

These drug, reducing bleeding episodes and treatment complications, offers a potential long-term solution or even a permanent cure, transforming the outlook for individuals with Hemophilia B. Table 3 explicitly shows research milestones have been made over the years since the first funding released for Hemophilia B in 1985.

**Table 3.** FDA-Approved treatments for Hemophilia B over time.

|  | Generic Name | Orphan Designation | Designation Date | Marketing Approval Date | Exclusivity End |
| --- | --- | --- | --- | --- | --- |
| 1 | Coagulation factor IX | Replacement treatment and prophylaxis of the hemorrhagic complications of hemophilia B. | 06/27/1989 | 08/20/1992 | 08/20/1999 |
| 2 | Coagulation Factor IX (human) | For use as replacement therapy in patients with hemophilia B for the prevention and control of bleeding episodes, and during surgery to correct defective hemostasis. | 07/05/1990 | 12/31/1990 | 12/31/1997 |
| 3 | Coagulation Factor IX (recombinant) | Treatment of hemophilia B | 10/03/1994 | 06/26/2020 |  |
| 4 | Coagulation Factor IX (recombinant) | Treatment of hemophilia B | 10/03/1994 | 10/05/2021 |  |
| 5 | Coagulation Factor IX (recombinant) | Treatment of hemophilia B | 10/03/1994 | 02/11/1997 | 02/11/2004 |
| 6 | Coagulation factor IX (recombinant), Fc fusion protein | Control and prevention of hemorrhagic episodes in patients with hemophilia B (congenital factor IX deficiency or Christmas disease) | 10/30/2008 | 03/28/2014 | 03/28/2021 |
| 7 | Coagulation Factor IX (Recombinant), GlycoPEGylated | Routine prophylactic administration for prevention of bleeding in patients with hemophilia B (Christmas disease) | 03/18/2013 | 07/29/2022 |  |
| 8 | Coagulation factor IX (recombinant); coagulation factor IX | Prophylactic use to prevent or reduce the frequency of bleeding episodes in patients with hemophilia B (routine prophylaxis in patients | 10/31/2012 | 09/12/2014 | 09/12/2021 |
|  | (recombinant) | where there is no evidence or suspicion of bleeding) |  |  |  |
| 9 | coagulation factor IX (recombinant);<br>coagulation factor IX (recombinant) | Prophylactic use to prevent or reduce the frequency of bleeding episodes in patients with hemophilia B (routine prophylaxis in patients where there is no evidence or suspicion of bleeding) | 10/31/2012 | 06/26/2013 | 06/26/2020 |
| 10 | Etranacogene dezaparvovec-drlb | Treatment of Hemophilia B | 04/17/2019 | 11/22/2022 | 11/22/2029 |
| 11 | Fidanacogene elaparvovec-dzkt | Treatment of hemophilia B | 09/21/2015 | 04/25/2024 | 04/25/2031 |
| 12 | Recombinant fusion protein linking coagulation factor IX with albumin (rIX-FP) | Treatment of patients with congenital factor IX deficiency (hemophilia B) | 04/27/2012 | 03/04/2016 | 03/04/2023 |

#### Cypher Query 3

MATCH (n:Project)-[r:RESEARCHED_BY]-(g:GARD)

MATCH (n)--(c:CoreProject)--(p:Publication)

WHERE g.GardId=‘GARD:0008732’ and r.semantic_similarity > 0.6

RETURN n

## 5. Discussion

In this work, we developed a NIH grant knowledge graph using Neo4j by integrating data on 2,067 GARD diseases linked to over 320,000 NIH funded projects, facilitating data-driven medical breakthroughs in rare diseases. In addition to the BiomedBERT model, we also evaluated other models, including BlueBert [33], BioMed-RoBERTa [34], biobert [35], and Bio_ClinicalBERT [36], for the semantic similarity task using basic examples. We observed that BiomedBERT outperformed the other models, prompting us to proceed with fine-tuning it. Through two detailed case studies, we showcased the utility of this knowledge graph in analyzing funding patterns within rare diseases, systematically identifying new research gaps and opportunities. Furthermore, it facilitates the dissemination of scientific findings and evidence to funding agencies, aiding in the informed distribution of research funds dedicated to rare diseases.

### 5.1. Scientific Observations from Disease Mapping

Considering the context surrounding rare disease names mentioned in the text, we introduced semantic similarity and a confidence score to prioritize the mapping results after we obtained mapped text based on exact string match. By combining both scores, we were able to determine whether the project’s research focus is on the identified rare disease. This section outlines how to effectively apply these scores to improve the mapping results for rare diseases. Applying the multiplication of the normalized confidence score and the semantic similarity score as a function, we achieved an accuracy of 0.77 in identifying projects focused on rare diseases (please refer to Table 1 in the PerfomanceResults.docx). When using the average of the two scores, we achieved an accuracy of 0.78 (please refer to Table 2 in the PerfomanceResults.docx,). In general, we can extend various functions with these two scores to reach higher performance on mapping, for instance, different normalization strategies, or different thresholds to be applied. We define a function in mathematical terms that incorporates different weights for two variables: the project normalized confidence score and the project semantic similarity score. We evaluate this function against specified thresholds; if the value exceeds the threshold, we consider the result relevant.

We performed a grid search for each goal, minimizing FP or maximizing accuracy, to identify the top three choices regarding score threshold, normalization method, and function (please refer to Table 3 in the PerfomanceResults.docx,). If FP is critical, one should consider only those with a confidence score of 1 and/or a semantic similarity above 0.9. However, if more related projects to investigate, lower those thresholds should be recommended.

### 5.2. Scientific Insights Derived from the Case Studies

We performed two case studies to demonstrate the use of rare disease related grant funding data for rare disease research. The funding landscape analysis not only provides a comprehensive view of the current landscape and highlighting areas for potential growth and collaboration in the rare disease area, but also allow researchers and specialists to identify potential collaborators and resourceful regions, explore and identify regional disparities in funding. Analyzing funding data on specific rare diseases provides a systematic approach to outlining a comprehensive research spectrum. It also generates scientific evidence to support rare disease research discoveries. In our second case study, we assessed research landscape analysis on Hemophilia B (GARD:0008732). This assessment can help scientists find a roadmap to discover the mechanisms of rare diseases systematically. Additionally, it becomes feasible to cluster research papers based on funded projects or similar research topics, enabling researchers to systematically track research pathways and programmatically prepare training data for future studies.

### 5.3. Future Work

In this paper, by taking context information into consideration while we mapped the projects to rare diseases, we measured the semantic similarity score by building our datasets to fine-tune the BioBERT model for this task.

We examined the Large Language Model (LLM) ChatGPT-3.5 [37] to detect rare diseases. While the LLM demonstrates proficiency in disease identification, it faces limitations in efficiently detecting rare diseases due to its training datasets needing more specificity in this domain. Additionally, ChatGPT-3.5 can provide varying responses to the same prompt in different runs, and it has limitations in accurately determining whether the primary focus of a text is on a rare disease. Further research on using other LLMs for this purpose should be explored.

As the next step, we plan to generate new datasets to retrain embedding models that cover various biomedical or biological categories, including ‘Disease or Syndrome,’ ‘Mental or Behavioral Dysfunction,’ ‘Amino Acid, Peptide, or Protein,’ and ‘Pharmacologic Substance. Although we applied the proposed framework to NIH funding data to identify rare disease-related projects, the framework can easily be adapted to other types of free text to enhance databases related to rare diseases. Looking ahead, we plan to improve our rare disease databases by extending our current NIH grant data with additional resources from institutions such as Stanford [38], Harvard [39], Duke Medicine [40], and the CZI Community Fund [41]. Due to the limited data and knowledge about rare diseases compared to common diseases, collaborative efforts are essential. We propose developing methods to enhance research collaboration by connecting principle investigators with the shared research interests through our funding knowledge graph. This approach will allow scientists to combine patients, data, experience, and resources, fostering more innovative research on rare diseases.

## 6. Conclusion

In conclusion, our paper addresses the formidable challenges of rare diseases in the medical and research domains, characterized by the limited and unstructured nature of available information. We presented a method to identify rare disease related text using two scores, namely confidence score and semantic similarity score and assess the text relevance to the identified rare diseases. We applied our methods to the NIH grant data from NIH RePORTER, which offers a rich resource for supporting rare disease research. Overall, the results and discussions affirm the potential of our algorithm to significantly enhance accuracy and reduce FP in rare disease identification within NIH grant database, contributing valuable insights to the field of rare disease research.

## Data Availability

All data produced in the present study are available upon reasonable request to the authors

## Acknowledgement

Not applicable.

## Contributions

JV: data collection, pipeline development, manual review of Set 1, and manuscript writing. SM and YX: manual review of the Set 1 and manuscript editing; QZ: study conception, manual review of the Set 1 and manuscript writing. All authors reviewed and approved the manuscript.

## Data availability

The data applied in this study can be accessed at https://rdas.ncats.nih.gov/browser/, and https://github.com/ncats/RDAS/tree/master/RDAS_GFKG/OrphanMap.

## Declarations

### Ethics approval and consent to participate

Not applicable.

### Consent for publication

Not applicable.

## Footnotes

1 RareDiseaseExcludingList.docx for more information.

2 For example, familial pyrimidinemia is a synonym of dihydropyrimidine dehydrogenase deficiency. We consider both familial pyrimidinemia and pyrimidinemia familial when checking through the text. We limit the bag-of-words approach to only two-word combinations to avoid complicating the rare disease word list. For instance, if the name is ‘hyperphalangy-clinodactyly of index finger with Pierre Robin syndrome’, applying the bag-of-words metho would result in 8! different combinations. Additionally, the probability of encountering these combinations rather than the original form is low.

3 For example, we disregard words like “td”, “ahc”, and “bpes”. The number five is chosen based on experience. Selecting the appropriate number involves a trade-off between false positives and false negatives.

1 The interactive version of Figure 5B can be accessed online via Tableau Public, featuring filters for institution names, grant funding ($), and research outputs: https://public.tableau.com/app/profile/jab.valinejad/viz/GrantFundingDistributionforRareDiseaseResearchAcrossU_S_Institutions/GrantFundingDistributionforRareDiseaseResearchAcrossU_S_Institutions

